# Ultra-early hematoma expansion is associated with ongoing hematoma growth and poor functional outcome

**DOI:** 10.1101/2024.12.02.24318363

**Authors:** Chloe A. Mutimer, Teddy Y. Wu, Henry Zhao, Leonid Churilov, Bruce C.V. Campbell, Andrew Cheung, Atte Meretoja, Timothy J. Kleinig, Philip M. Choi, Henry Ma, Geoffrey C. Cloud, Rohan Grimley, Darshan Shah, Annemarei Ranta, Karim Mahawish, Vignan Yogendrakumar, Gagan Sharma, Geoffrey A. Donnan, Stephen M. Davis, Nawaf Yassi, the STOP-MSU Trial Investigators

## Abstract

**INTRODUCTION:** There are limited data on ultra-early hematoma growth dynamics and their clinical impact in primary intracerebral hemorrhage (ICH). We aimed to (a) estimate the incidence of hematoma expansion within the hyperacute period of ICH, (b) describe hematoma dynamics over time, (c) investigate the associations between ultra-early hematoma expansion and clinical outcomes after ICH, and (d) assess the effect of tranexamic acid on ultra-early hematoma expansion.

**METHODS:** We performed a planned secondary analysis of the STOP-MSU international multicenter randomized controlled trial. The trial compared tranexamic acid with placebo in 201 patients with primary ICH presenting within 2 hours of symptom onset. Repeat CT imaging ∼1 hour after treatment commencement was encouraged.

Patients who underwent re-imaging up to 3 hours from baseline imaging were included in this descriptive study. Hematoma expansion was defined as either a ≥33% or ≥6 ml increase from baseline hematoma volume.

**RESULTS:** We included 105 of the 201 patients who had 1-hour imaging (median age 66 years, 40% female, 53% tranexamic acid). Median time from onset to baseline imaging was 74min (IQR 56-87 min), and between baseline and 1-hour imaging was 95 min (IQR 74-132 min). Forty-one patients (39%) had ultra-early hematoma expansion. These patients had larger baseline hematoma volumes (15.9 ml vs 9.1 ml, p=0.03) compared to those with no early hematoma expansion.

Hematoma growth rate declined over time compared to the onset-to-baseline imaging period (clustered median regression p<0.01). In 92 patients with both 1-hour and 24-hour re-imaging, 9/31 (29%) of those with ultra-early hematoma expansion had further expansion between the 1-hour and 24-hour scan, compared to only 4/61(6.6%) without ultra-early expansion (p<0.01). Of those 61 patients, there were 10 (16.4%) who fulfilled the expansion definition by 24 hours.

Ultra-early hematoma expansion was associated with poor functional outcomes (mRS 3-6; aOR 3.87 [1.21-12.40], p=0.02) and mortality (aOR 6.16 [95% CI 2.15-17.68], p<0.01), adjusted for treatment group.

There was no observed effect of tranexamic acid treatment on ultra-early hematoma expansion (41% vs. 37%, p=0.65).

**CONCLUSIONS:** Most hematoma growth occurs in the ultra-early period. The presence of hyperacute hematoma expansion is associated with ongoing hematoma growth, poor functional outcomes and mortality, and represents a target for therapeutic intervention.

## Introduction

Hematoma expansion after initial imaging occurs in up to one third of patients with primary intracerebral hemorrhage within the first 24 hours and is strongly associated with neurological deterioration, poor functional outcome and mortality.^1,2^ The attenuation of hematoma expansion has been identified as one of the primary therapeutic targets in intracerebral hemorrhage.^3^ A large systematic review has shown that most hematoma growth occurs within the first 6 hours.^4^ The dynamics of hematoma growth in this timeframe are not well understood and there are very limited data in the sub-3 hour timeframe, with only three other small studies.^5-7^

To further understand hematoma growth in the hyperacute timeframe, we performed a pre-planned secondary analysis of the STOP-MSU international randomized controlled trial and assessed hematoma characteristics by obtaining repeat computed tomography (CT) imaging 1-hour after baseline imaging. We aimed to (a) estimate the incidence of hematoma expansion within the ultra-early period of ICH, (b) describe hematoma dynamics over time, with a specific focus on the ultra-early period, (c) investigate the associations between ultra-early hematoma expansion and clinical outcomes after ICH, and (d) assess the effect of tranexamic acid on ultra-early hematoma expansion.

## Methods

### Study design and participants

This study is a pre-planned secondary analysis of the STOP-MSU (2018-2023) international multicenter randomized controlled trial in primary intracerebral hemorrhage.^8^ Briefly, the STOP-MSU trial compared intravenous tranexamic acid (1 g over 10 minutes, followed by 1 g over 8 hours) with placebo (saline, matched dosing regimen) in patients presenting within 2 hours of symptom onset.

Pre-treatment and 24-hour CT imaging was performed as part of the prespecified imaging protocol. Hyperacute repeat imaging at ∼1 hour from treatment was performed as part of an optional substudy in a proportion of participants. Only patients who had “1-hour imaging” (defined as ≤3 hours from baseline scan) were included in the present study.

### Standard protocol approvals, registrations, and patient consents

The STOP-MSU study protocol was approved by the respective ethics committees of the recruiting sites. Written informed consent was obtained from the participant or a legal representative before enrolment according to the Declaration of Helsinki, although emergency treatment followed by consent for continued participation was allowed in some jurisdictions.

### Data sharing

Deidentified data will be made available on reasonable request by email communication to the corresponding author following review and approval of a research proposal by the trial executive committee, with a signed data access agreement.

### Neuroimaging analysis

Hematoma volume (intracerebral and intraventricular) was measured using non-contrast CT and a validated semi-automated planimetric method,^9^ with baseline and 24-hour imaging assessed as part of the STOP-MSU study at the central imaging lab (Christchurch, New Zealand). In the protocol, hyperacute imaging at 1 hour after treatment initiation was encouraged, but not mandated. Hematoma volumes from ultra-early re-imaging were measured using the same method by two authors (CM and TW). Both investigators were blinded to treatment outcomes.

For the purpose of investigating hematoma growth dynamics, we divided each participant’s imaging analyses into the following time epochs: *<u>baseline</u>* – the period from onset to baseline imaging; *<u>ultra-early</u>*: the period from baseline imaging to 1-hour imaging; and *<u>interval</u>*: the period from 1-hour to 24 hour re-imaging.

### Outcomes

The primary outcome was hematoma expansion on 1-hour imaging compared to baseline, with hematoma expansion defined as an increase of ≥6 ml or ≥33% of hematoma volume (referred to as “ultra-early hematoma expansion).^10^ Interval hematoma expansion was defined by the same definition occurring between the 1-hour and 24-hour scans, and hematoma expansion at 24-hours was that occurring between the baseline and 24-hour scans. This was the same definition used in the primary study. Hematoma growth rates were defined as hematoma volume divided by time in hours from symptom onset for baseline growth rate; or change in hematoma volume divided by time in hours from previous scan for 1-hour and interval growth rates.^11^

### Statistical analysis

We compared baseline features and radiological and clinical outcomes of participants using Fisher’s exact test for categorical outcomes, and Mann-Whitney U test for continuous outcome as appropriate. This was used to compare (1) participants with and without 1-hour imaging, (2) participants with and without ultra-early hematoma expansion and (3) participants by randomisation group (placebo or tranexamic acid). We tested whether ultra-early hematoma expansion was associated with functional outcomes on univariate logistic regression. This study is of a descriptive nature, and the sample size is not large enough to adjust for covariates, therefore adjustment of odds ratios was not performed.^12,13^ To address potential bias introduced by more clinically severe patients being more likely to have ultra-early re-imaging, we performed a sensitivity analysis excluding patients who had significant early clinical deterioration. To investigate the association of ultra-early and interval hematoma expansion with 24-hour expansion, receiver operating characteristic analysis was performed with the outcome variable being the presence or absence of 24-hour hematoma expansion.

Statistical analyses were performed with STATA-SE version 18, and 2-sided p-values ≤0.05 were considered statistically significant.

## Results

### Baseline characteristics and ultra-early hematoma expansion

Of the 201 patients enrolled into the STOP-MSU study, 105 (52.2%) had baseline and ultra-early re-imaging, and 92 (45.8%) had baseline, ultra-early and 24-hour imaging. The majority of patients with ultra-early re-imaging (96.2%) were recruited in Australia or New Zealand, and 32.3% were recruited from the mobile stroke unit (Supplementary Table S1).

For participants with baseline and 1-hour imaging, the median age was 66 years (IQR 55-77 years), 40% were female and 30.5% were on an antiplatelet agent. The median time from onset to baseline imaging was 74 minutes (IQR 56-87 minutes), and between baseline and 1-hour imaging was 95 minutes (IQR 74-132 minutes).

Median time from symptom onset to 1-hour imaging was 174 minutes (IQR 148.8-199.8 minutes). Forty-one (39.0%) of patients had hematoma expansion on 1-hour imaging. Baseline demographic and clinical characteristics were similar between participants with ultra-early hematoma expansion and those without, with the exception of higher NIHSS and lower GCS in the ultra-early hematoma expansion group (not statistically significant) (Table 1). Patients with ultra-early hematoma expansion had higher baseline hematoma volumes (15.9 ml vs. 9.1 ml, p=0.032), and ultra-early hematoma expansion was numerically more frequent in patients with lobar compared to deep intracerebral hemorrhage (52% vs. 35%, p=0.128).

**Table 1:**
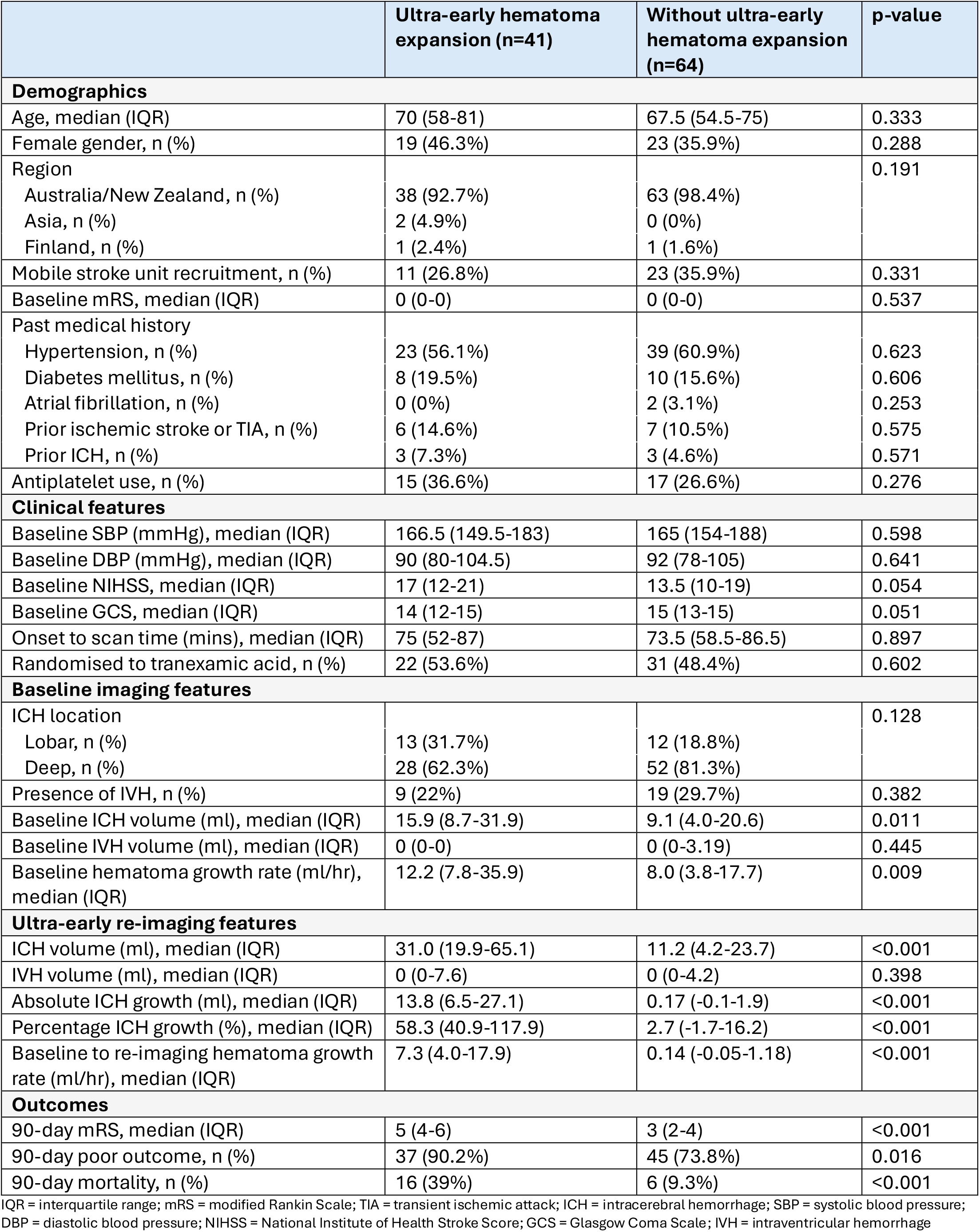
Characteristics of participants with and without ultra-early hematoma expansion.

|  | Ultra-early hematoma expansion (n=41) | Without ultra-early hematoma expansion (n=64) | p-value |
| --- | --- | --- | --- |
| **Demographics** |  |  |  |
| Age, median (IQR) | 70 (58-81) | 67.5 (54.5-75) | 0.333 |
| Female gender, n (%) | 19 (46.3%) | 23 (35.9%) | 0.288 |
| Region |  |  | 0.191 |
| Australia/New Zealand, n (%) | 38 (92.7%) | 63 (98.4%) |  |
| Asia, n (%) | 2 (4.9%) | 0 (0%) |  |
| Finland, n (%) | 1 (2.4%) | 1 (1.6%) |  |
| Mobile stroke unit recruitment, n (%) | 11 (26.8%) | 23 (35.9%) | 0.331 |
| Baseline mRS, median (IQR) | 0 (0-0) | 0 (0-0) | 0.537 |
| Past medical history |  |  |  |
| Hypertension, n (%) | 23 (56.1%) | 39 (60.9%) | 0.623 |
| Diabetes mellitus, n (%) | 8 (19.5%) | 10 (15.6%) | 0.606 |
| Atrial fibrillation, n (%) | 0 (0%) | 2 (3.1%) | 0.253 |
| Prior ischemic stroke or TIA, n (%) | 6 (14.6%) | 7 (10.5%) | 0.575 |
| Prior ICH, n (%) | 3 (7.3%) | 3 (4.6%) | 0.571 |
| Antiplatelet use, n (%) | 15 (36.6%) | 17 (26.6%) | 0.276 |
| **Clinical features** |  |  |  |
| Baseline SBP (mmHg), median (IQR) | 166.5 (149.5-183) | 165 (154-188) | 0.598 |
| Baseline DBP (mmHg), median (IQR) | 90 (80-104.5) | 92 (78-105) | 0.641 |
| Baseline NIHSS, median (IQR) | 17 (12-21) | 13.5 (10-19) | 0.054 |
| Baseline GCS, median (IQR) | 14 (12-15) | 15 (13-15) | 0.051 |
| Onset to scan time (mins), median (IQR) | 75 (52-87) | 73.5 (58.5-86.5) | 0.897 |
| Randomised to tranexamic acid, n (%) | 22 (53.6%) | 31 (48.4%) | 0.602 |
| **Baseline imaging features** |  |  |  |
| ICH location |  |  | 0.128 |
| Lobar, n (%) | 13 (31.7%) | 12 (18.8%) |  |
| Deep, n (%) | 28 (62.3%) | 52 (81.3%) |  |
| Presence of IVH, n (%) | 9 (22%) | 19 (29.7%) | 0.382 |
| Baseline ICH volume (ml), median (IQR) | 15.9 (8.7-31.9) | 9.1 (4.0-20.6) | 0.011 |
| Baseline IVH volume (ml), median (IQR) | 0 (0-0) | 0 (0-3.19) | 0.445 |
| Baseline hematoma growth rate (ml/hr), median (IQR) | 12.2 (7.8-35.9) | 8.0 (3.8-17.7) | 0.009 |
| **Ultra-early re-imaging features** |  |  |  |
| ICH volume (ml), median (IQR) | 31.0 (19.9-65.1) | 11.2 (4.2-23.7) | <0.001 |
| IVH volume (ml), median (IQR) | 0 (0-7.6) | 0 (0-4.2) | 0.398 |
| Absolute ICH growth (ml), median (IQR) | 13.8 (6.5-27.1) | 0.17 (-0.1-1.9) | <0.001 |
| Percentage ICH growth (%), median (IQR) | 58.3 (40.9-117.9) | 2.7 (-1.7-16.2) | <0.001 |
| Baseline to re-imaging hematoma growth rate (ml/hr), median (IQR) | 7.3 (4.0-17.9) | 0.14 (-0.05-1.18) | <0.001 |
| **Outcomes** |  |  |  |
| 90-day mRS, median (IQR) | 5 (4-6) | 3 (2-4) | <0.001 |
| 90-day poor outcome, n (%) | 37 (90.2%) | 45 (73.8%) | 0.016 |
| 90-day mortality, n (%) | 16 (39%) | 6 (9.3%) | <0.001 |

IQR = interquartile range; mRS = modified Rankin Scale; TIA = transient ischemic attack; ICH = intracerebral hemorrhage; SBP = systolic blood pressure; DBP = diastolic blood pressure; NIHSS = National Institute of Health Stroke Score; GCS = Glasgow Coma Scale; IVH = intraventricular hemorrhage

### Hematoma growth rates

The evolution of individual patient hematoma volumes over time is shown in Figure 1. Median intracerebral and intraventricular hemorrhage volumes across the time epochs are summarised in Table 2. The median baseline hematoma growth rate for all patients was 10.3 ml/hr (IQR 4.34-26.82 ml/hr), the median ultra-early hematoma growth rate was 1.34 ml/hr (IQR 0.06-5.51 ml/hr), and the median interval hematoma growth rate was 0.1 ml/hr (IQR -0.04-0.09 ml/hr). Median hematoma growth rate in the ultra-early and interval time epochs was significantly lower than the onset-to-baseline imaging period (clustered median regression p<0.01) (Figure 2). There were 22/105 (21%) cases of hematoma regression/retraction between baseline and 1-hour imaging compared with 39/92 (42.4%) cases of hematoma regression between 1-hour and 24 hour imaging.

**Table 2:** Hematoma volumes for participants with ultra-early reimaging (median, IQR)

|  | ICH volume (ml) | IVH volume (ml) |
| --- | --- | --- |
| Baseline (n=105) | 12.42 (5.71-23.57) | 0 (0-0.65) |
| Ultra-early scan (n=105) | 17.87 (6.76-33.76) | 0 (0-5.18) |
| 24-hour scan (n=92) | 15.28 (6.8-33.62) | 0 (0-3.33) |

IQR = interquartile range; ICH = intracerebral hemorrhage; IVH = intraventricular hemorrhage

**Figure 1:**
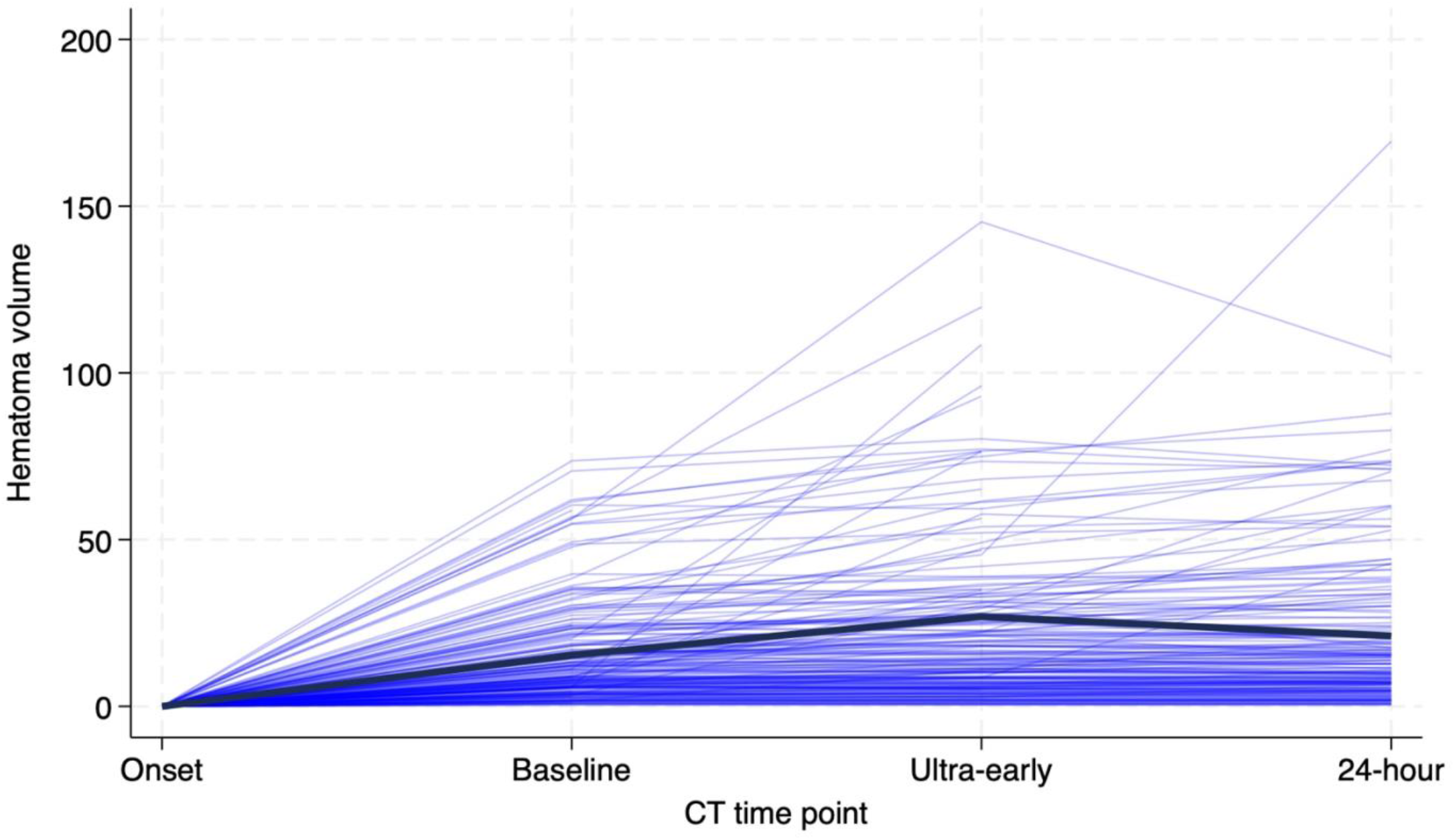
spaghetti plot of hematoma volume over the 24-hour time period as measured on non-contrast CT for all patients. Bold line is the population mean across the time points.

**Figure 2:**
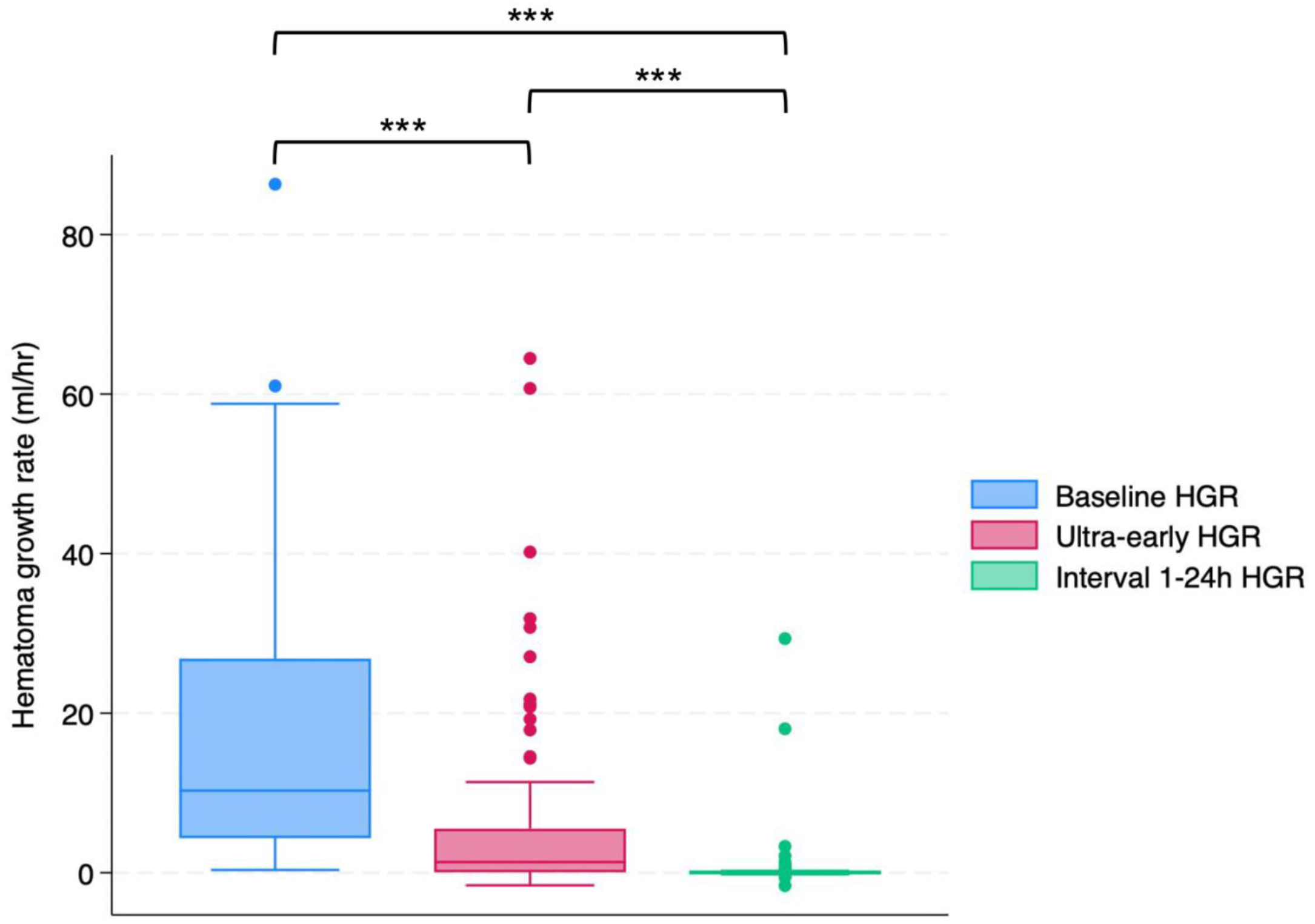
Box plot for hematoma growth rate across the three timepoints, from onset to baseline imaging (baseline HGR), baseline to ultra-early imaging (ultra-early HGR) and ultra-early imaging to 24-hour imaging. HGR = hematoma growth rate.

### 24-hour imaging characteristics

Follow-up imaging features of the 92/105 (87.6%) patients with scans at baseline, 1-hour and 24-hours are shown in Table 3 Of the 13 patients without 24-hour imaging available, 8 died (61.5%), 3 had operative intervention and 2 did not have repeat imaging. The median time from baseline to 24-hour imaging was 25.1 hours (IQR 23.3-26.4 hours). The median absolute hematoma volume change between the ultra-early and 24-hour scans was 0.16ml (IQR -1.04-2.08ml), median hematoma volume change percentage was 2.6% (IQR-8.6-12.8%) and median growth rate was 0.01ml/hr (IQR -0.04-0.24ml/hr). Patients with ultra-early hematoma expansion were more likely to have additional interval hematoma expansion at 24-hours than those who did not have ultra-early growth (73.0% vs. 7.3%, p<0.001). The sensitivity of ultra-early hematoma expansion for 24-hour hematoma expansion was 0.73 (95%CI 0.56-0.86), with a specificity of 0.94 (95%CI 0.82-0.98), a positive predictive value of 0.87 (95%CI 0.72-0.95) and a negative predictive value of 0.84 (95%CI 0.75-0.90). The area under the receiver operator curve was 0.83 (95%CI 0.75-0.91).

**Table 3:** Follow-up imaging features of patients with ultra-early scans (n=92)

|  | Ultra-early hematoma expansion (n=31) | Without ultra-early hematoma expansion (n=61) | p-value |
| --- | --- | --- | --- |
| **Interval time frame (1 to 24-hours)** |  |  |  |
| Interval absolute ICH growth (ml), median (IQR) | -0.1 (-2.2-6.3) | 0.2 (-0.7-1.5) | 0.715 |
| Interval percentage ICH growth (%), median (IQR) | -0.7 (-10.3-15.9) | 3.4 (-8.6-12.3) | 0.782 |
| Interval hematoma growth rate (ml/hr), median (IQR) | -0.01 (-0.1-0.2) | 0.01 (-0.03-0.07) | 0.630 |
| Interval hematoma expansion*, n (%) | 9 (29.0%) | 4 (6.6%) | 0.004 |
| **24-hour imaging features** |  |  |  |
| 24-hour ICH volume (ml), median (IQR) | 28.8 (17.0-60.2) | 9.8 (4.9-22.8) | <0.001 |
| 24-hour IVH volume (ml), median (IQR) | 0 (0-4.3) | 0 (0-2.9) | 0.240 |
| Hematoma expansion at 24-hours, n (%) | 27 (87.1%) | 4 (6.6%) | <0.001 |

\*Significant hematoma expansion between ultra-early and 24-hour scans. IQR = interquartile range; ICH = intracerebral hemorrhage; IVH = intraventricular hemorrhage

There were 13/92 patients (14.1%) with interval hematoma expansion between ultra-early and 24-hour imaging, which was more likely in patients who already had ultra-early hematoma expansion (29.0% vs. 6.6%, p=0.004). Of the 61 patients that did not have ultra-early hematoma expansion, 10 (16.4%) met the definition for hematoma expansion at 24-hours. Clinical and imaging characteristics of these patients are shown in Supplementary Table S3. The sensitivity of ultra-early hematoma expansion for interval hematoma expansion (between ultra-early imaging and 24-hour imaging) was 0.69 (95%CI 0.39-0.91), with a specificity of 0.72 (95%CI 0.61-0.82), a positive predictive value of 0.55 (95%CI 0.43-0.68) and a negative predictive value of 0.82 (95%CI 0.67-0.91). The area under the receiver operator curve was 0.70 (95%CI 0.56-0.84).

### Clinical outcomes

Patients in the ultra-early hematoma expansion group were more functionally impaired at 90 days (median mRS 5 [IQR 4-6] vs. 3 [IQR 2-4], p<0.001) and experienced higher mortality. Ordinal mRS analysis is shown in Figure 3. In univariable logistic regression analysis, ultra-early hematoma expansion was associated with 90-day poor functional outcome and 90-day mortality (Table 4).

**Table 4:**
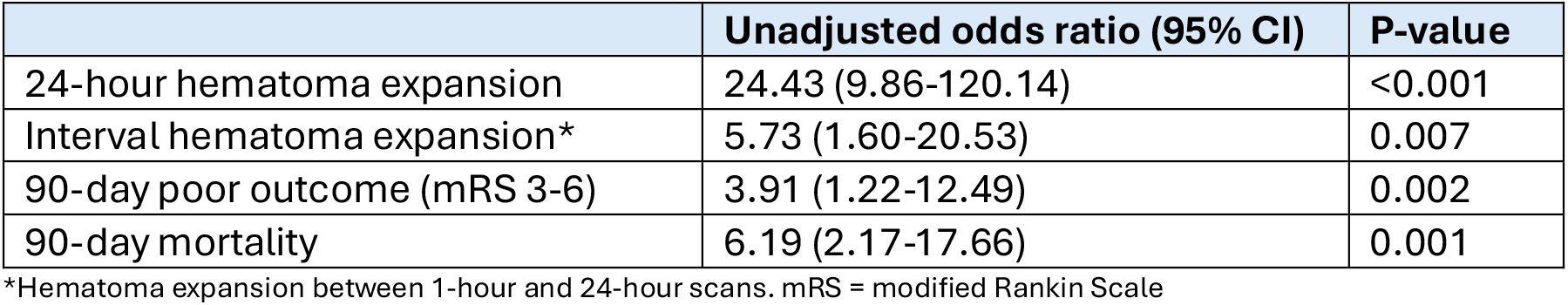
Univariate logistic regression for ultra-early hematoma expansion and association with outcomes.

**Figure 3:**
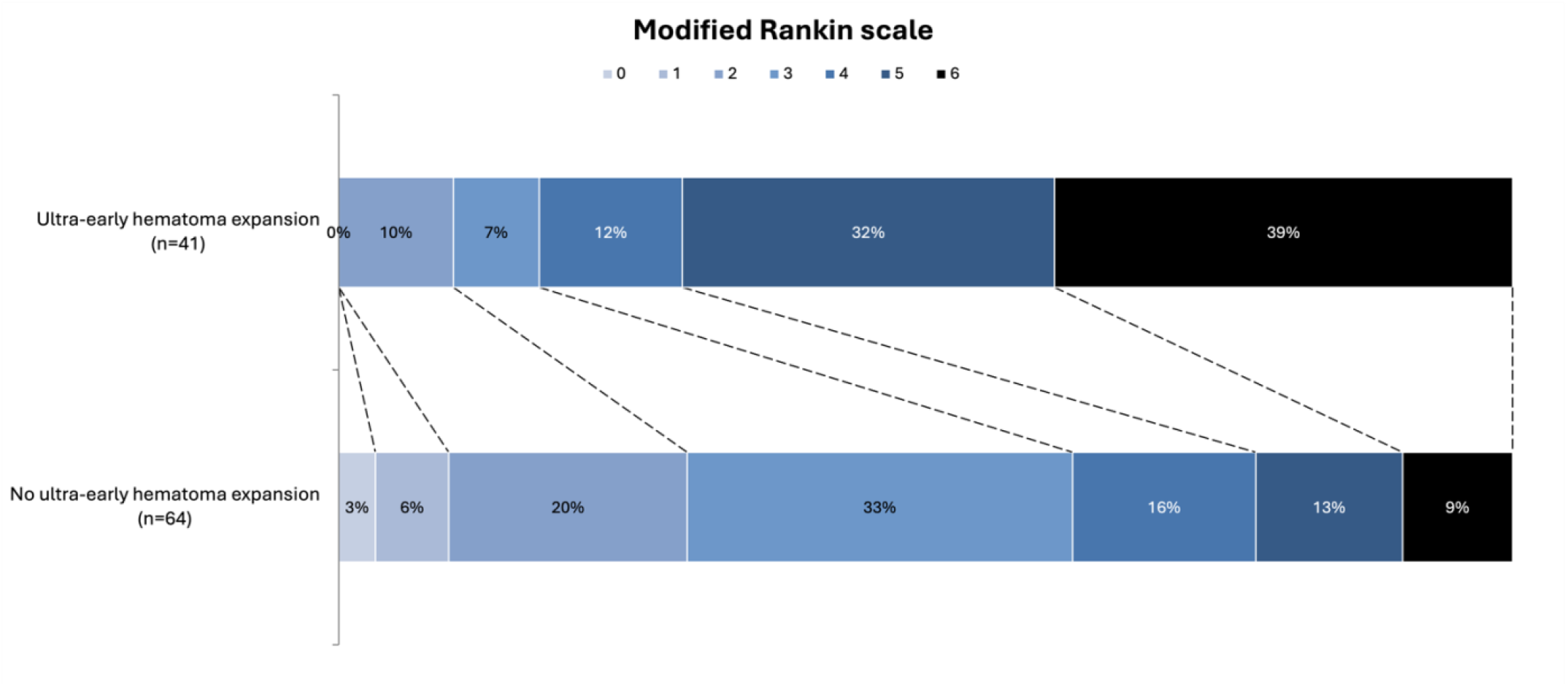
Modified Rankin Scale scores at 90 days for participants with ultra-early scans (unadjusted OR 1.97 [95% CI 1.18-2.76], p<0.001)

Of the 105 patients with 1-hour imaging, 11 (10.5%) had early deterioration reported as a serious adverse event. Sensitivity analysis excluding these patients is shown in Supplementary Table S2, and ultra-early hematoma expansion remained associated with 24-hour hematoma expansion, 90-day poor functional outcome and 90-day mortality.

### Effect of tranexamic acid

Baseline characteristics for included patients randomised to either tranexamic acid or placebo were similar (Supplementary Table S4). There was no significant difference in rates of ultra-early hematoma expansion between tranexamic acid and placebo groups (41.1% vs. 36.7%, p=0.650) (Table 5). There was also no difference between absolute and relative (percentage) intracerebral hemorrhage growth at 1-hour between groups. Ordinal mRS analysis is shown in Figure S1. Of the 92 patients with 24-hour imaging available for review, 49 (53.2%) were randomised to tranexamic acid and overall, 24-hour hematoma volumes were larger in the tranexamic acid group (median 18.6ml vs. 13.7ml, p=0.025). However, despite this difference in 24-hour volume there was no significant difference in hematoma expansion at 24 hours compared with baseline between groups (42.9% in the tranexamic acid group vs. 37.9% in the control group, p=0.581).

**Table 5:** Imaging features and outcomes of patients with ultra-early scans by randomisation group.

|  | Tranexamic acid (n=56) | Placebo (n=49) | p-value |
| --- | --- | --- | --- |
| **Baseline imaging features** |  |  |  |
| ICH location |  |  | 0.257 |
| Lobar, n (%) | 16 (28.6%) | 9 (18.4%) |  |
| Deep, n (%) | 40 (71.4%) | 40 (81.6%) |  |
| Presence of IVH, n (%) | 14 (25%) | 14 (28.6%) | 0.633 |
| Baseline ICH volume (ml), median (IQR) | 13.3 (5.8-30.3) | 12.3 (5.5-20.3) | 0.330 |
| Baseline IVH volume (ml), median (IQR) | 0 (0-0.7) | 0 (0-0.9) | 0.830 |
| Baseline hematoma growth rate (ml/hr), median (IQR) | 10.6 (4.7-31.8) | 8.3 (4.3-22.1) | 0.319 |
| **1-hour imaging features** |  |  |  |
| ICH volume (ml), median (IQR) | 21.9 (8.0-50.6) | 12.8 (5.5-27.5) | 0.088 |
| IVH volume (ml), median (IQR) | 0 (0-6.0) | 0 (0-5.0) | 0.847 |
| Absolute ICH growth (ml), median (IQR) | 2.6 (0.2-13.5) | 2.1 (0.01-7.8) | 0.489 |
| Percentage ICH growth (%), median (IQR) | 18.3 (1.9-46.0) | 17.8 (0.5-54.6) | 0.766 |
| Ultra-early hematoma expansion, n (%) | 23 (41.1%) | 18 (36.7%) | 0.650 |
| **90-day outcomes** |  |  |  |
| 90-day mRS, median (IQR) | 4 (3-5.5) | 3 (3-5) | 0.117 |
| 90-day poor outcome, n (%) | 45 (80.4%) | 37 (75.6%) | 0.549 |
| 90-day mortality, n (%) | 14 (25%) | 8 (16.3%) | 0.276 |

ICH = intracerebral hemorrhage; IVH = intraventricular hemorrhage; IQR = interquartile range; mRS = modified Rankin Scale

## Discussion

In this prespecified secondary analysis of the STOP-MSU trial, we found that most hematoma growth occurred in the ultra-early period of intracerebral hemorrhage. In our study, this corresponded to a median onset to re-imaging time of 174 minutes. This finding is supportive of the findings of larger in-hospital studies in which hematoma growth occurred mostly in a non-linear manner in the first 24-hours, with increased probability of expansion in patients presenting within 6 hours of symptom onset.^4^

We found that ultra-early hematoma expansion occurred in 39% of patients presenting within 2 hours of symptom onset. This is the first study to perform repeat imaging to assess hematoma growth for both in-hospital and mobile stroke unit patients. Our rates of ultra-early hematoma expansion are higher than those previously reported. In a study by Bowry et al. of 49 mobile stroke unit patients presenting in the ultra-early timeframe, 28% and 17% of patients presenting within 1 hour and 1-2 hours of symptom onset respectively had early hematoma expansion.^5^ Al-Ajlan et al. performed a planned secondary analysis of the SPOTLIGHT trial with ultra-early repeat imaging in 44 patients (rFVIIa versus placebo within 6 hours of onset in primary intracerebral haemorrhage with a spot sign).^7^ They too showed that absolute hematoma growth occurs between the onset of symptoms and ultra-early re-imaging. This is despite including patients in a later timeframe (up to 6 hours), with no stratification for patients presenting under 2 hours. There is one other study of repeat imaging in the mobile stroke unit setting of 17 intracerebral hemorrhage patients, of which 9 patients were imaged within 2 hours of symptom onset with an 11% rate of early hematoma expansion.^6^ However, alternate definitions of hematoma expansion were used in all papers, impacting the generalisability of these results.

The pathophysiology of hematoma expansion is complex. It has been proposed that the initial hematoma is formed due to a single vessel rupture, with rate of initial expansion dependent on the size of the vessel.^1^ There may then be shearing of other vessels resulting in secondary expansion.^14^ Interventions to limit hematoma expansion should therefore focus on patients presenting within this early timeframe, as currently being conducted in the TICH-3 trial (tranexamic acid within 4.5 hours of symptom onset) and the FASTEST trial (factor VIIa within 2 hours).^15,16^ This finding also supports the increasing shift to adopting time metrics for intracerebral hemorrhage-specific treatments such as acute blood pressure lowering and anticoagulation reversal within an hour of patient presentation, in a similar paradigm to ischemic stroke, to prevent further hematoma expansion.^17^

The optimal measure of hematoma growth is uncertain, with various definitions of hematoma expansion used in the literature. Definitions may take into account either relative growth (usually ≥33%) or absolute growth, which ranges from ≥6 to ≥12.5 ml, when comparing imaging at the 24-hour timeframe to baseline imaging.^10^ We used the definition of growth by either ≥6 ml or ≥33% from baseline imaging in keeping with the primary study, acknowledging our earlier timeframe. In our study, patients with ultra-early hematoma expansion had larger baseline hemorrhage volumes, worse functional outcomes and higher mortality compared to those without hematoma expansion. This is despite similar ages, rates of antiplatelet use, and rates of baseline intraventricular haemorrhage, all factors associated with poor prognosis in intracerebral hemorrhage.^4,18^ This suggests that this definition is still able to detect clinically significant hematoma expansion despite the ultra-early time period.

Minimally-invasive surgery for intracerebral hemorrhage is an emerging treatment option, with the recently published ENRICH trial showing benefit, although restricted to lobar hemorrhage.^19^ Minimally-invasive surgery can be considered for patients presenting outside of the ultra-early timeframe, with the inclusion of patients up to 24-hours from symptom onset, as shown by the median time from last known well time to surgery of 16.75 hours in ENRICH. There is therefore a need to understand which patients may continue to expand after the 3-hour timepoint, and thus may benefit from minimally-invasive surgery. In our study, ultra-early hematoma expansion was associated with interval hematoma expansion at the 24-hour timepoint, a finding that differs from the literature.^5^ This may be due to chance, or alternatively the quoted study may have been underpowered to detect a difference (n=8 with 1-hour hematoma expansion and follow-up imaging). Additionally, our study population which included in-hospital patients in addition to those from a mobile stroke unit may have unique hematoma growth dynamics given overall faster onset to imaging times. We had 10 patients who did not have ultra-early hematoma expansion with subsequent hematoma expansion at 24-hours when compared to baseline imaging. These patients were older, had higher rates of antiplatelet use and larger baseline hemorrhage volumes. Given the relatively small numbers, further studies are required to support these findings.

We found that hematoma expansion beyond the 1-hour imaging timepoint was rare in those without initial hematoma expansion, with retraction of hematoma volume seen at 24-hours compared to the ultra-early hematoma volume, with smaller median hematoma volumes at 24-hours compared to 1-hour imaging. Hematoma retraction was also found in a subset of participants in the primary STOP-MSU study.^8^ Animal models of intracerebral hemorrhage have shown that clot retraction can occur within a few hours of hematoma development, with shift of serum from the hematoma into surrounding tissue.^20^ Given hematoma retraction occurs within the first few hours of intracerebral hemorrhage onset, ultra-early hematoma expansion may be a more clinically meaningful treatment target or a clinical trial outcome for therapies seeking to minimise early hematoma expansion. This is further supported by the associations in this study with functional outcome and mortality.

In keeping with the findings of the primary analysis, which showed neutral effect of tranexamic acid on 24-hour hematoma expansion, tranexamic acid had no effect on ultra-early hematoma expansion. This secondary study is the first study to explore the effect of tranexamic acid in this early timeframe. Patients randomised to tranexamic acid had numerically larger intracerebral hemorrhage volumes on 1-hour and 24-hour imaging compared to the placebo group, suggesting that drivers for ultra-early hematoma growth may negate any effect of antifibrinolytic therapy in prevention of early hematoma expansion.

Strengths of this study include inclusion of patients recruited within the ultra-early timeframe, 2 hours from symptom onset, from emergency department presentations and mobile stroke units, allowing generalisability to both pre-hospital and in-hospital patients. This was partly achievable by the utilisation of a mobile stroke unit. Despite the small sample size, this study is the largest to date to report hematoma expansion after repeat ultra-early imaging, with only two other studies with repeat ultra-early imaging (median time from baseline to “1-hour imaging” of 2.3 hours).^5^ This study also contributes to the limited literature regarding intracerebral hemorrhage patients imaged within the first hour of symptom onset. Another strength is the utilisation of a central imaging lab for imaging analysis, with all volumes measured by semi-automated techniques.

Limitations of this study include its small sample size and it being a secondary analysis of a randomised controlled trial. Second, not all patients in the STOP-MSU study had ultra-early imaging, as it was an optional part of the protocol. This may have selected for patients at higher risk of hematoma expansion, with higher baseline hematoma volumes, higher rates of antiplatelet use and worse functional outcomes in patients who had ultra-early reimaging. However, median 24-hour hematoma volumes between groups were similar (Supplementary Table S1). We attempted to control for this potential bias by performing a sensitivity analysis excluding patients with early deterioration, with similar associations with poor radiological and clinical outcomes seen. Despite being a secondary analysis of an international study, the vast majority of patients included were from Australia or New Zealand potentially limiting generalisability. Additionally, blood pressure readings aside from baseline measurement were not available. Blood pressure control pre-hospital and in the early timeframe has recently been shown to be of functional benefit in intracerebral hemorrhage, in a patient population without baseline imaging.^21^ Finally, patients on anticoagulation were not included, so these findings cannot be extrapolated to anticoagulation-associated intracerebral hemorrhage.

In conclusion, we found that in patients with intracerebral hemorrhage presenting within 2 hours of onset, most hematoma growth occurs within the first 3 hours from symptoms onset, supporting previous data. Ultra-early hematoma expansion is associated with ongoing hematoma expansion at 24-hours, although interval growth was uncommon. Ultra-early hematoma expansion is also associated with poor clinical outcomes and should be a key therapeutic target in future clinical trials.

## Abbreviations

CT: Computed tomography
DBP: Diastolic blood pressure
GCS: Glasgow coma scale
ICH: Intracerebral hemorrhage
IQR: Interquartile range
IVH: Intraventricular hemorrhage
mRS: Modified Rankin scale
NIHSS: National Institutes of Health Stroke Scale
SBP: Systolic blood pressure
TIA: Transient ischemic attack

## Acknowledgments

STOP-MSU Investigators*:

Helen Dewey (Box Hill Hospital, Australia); Helen Brown, Emma Harrison and Michael Devlin (Princess Alexandra Hospital, Australia); Neil Spratt (John Hunter Hospital, Australia); Vincent Thijs (Austin Hospital, Australia); Lauren Sanders (St Vincent’s Hospital Melbourne, Australia); John Worthington (Royal Prince Alfred Hospital, Australia); Ben Clissold (Geelong University Hospital, Australia); Jiann-Shing Jeng (National Taiwan University Hospital, Taiwan); Hao-Kuang Wang (E-DA Hospital, Taiwan); Duy Ton Mai (Bach Mai Hospital, Viet Nam); Dang Phuc Duc (Military 103 Hospital, Viet Nam); and Nguyen Thai My Phuong and Nguyen Thi Huong (Nguyen Tri Phuong Hospital, Viet Nam).

Data Safety Monitoring Board Committee (STOP-MSU):

Barry Snow (chair) Richard Macdonell, John Macdonell, John King, Richard Stark, Neil Anderson, John Attia, Monique Kilkenny and John Kolbe.

## Supplementary material

**Supplementary Table S1:** Baseline characteristics of participants with and without ultra-early scans.

|  | With 1 hour imaging<br>(n=105) | Without 1 hour<br>imaging (n=96) | p-value |
| --- | --- | --- | --- |
| **Demographics** |  |  |  |
| Age, median (IQR) | 70 (57-78) | 64 (54-76) | 0.117 |
| Female gender, n (%) | 42 (40%) | 40 (42.7%) | 0.762 |
| Region |  |  | <0.001 |
| Australia/New Zealand, n (%) | 101 (96.2%) | 44 (45.8%) |  |
| Asia, n (%) | 2 (1.90%) | 36 (37.5%) |  |
| Finland, n (%) | 2 (1.90%) | 16 (16.6%) |  |
| Mobile stroke unit recruitment, n (%) | 34 (32.3%) | 10 (10.4%) | <0.001 |
| Baseline mRS, median (IQR) | 0 (0-0) | 0 (0-0) | 0.539 |
| Past medical history |  |  |  |
| Hypertension, n (%) | 62 (59.0%) | 66 (68.8%) | 0.153 |
| Diabetes mellitus, n (%) | 18 (17.1%) | 18 (18.8%) | 0.767 |
| Atrial fibrillation, n (%) | 2 (1.90%) | 5 (5.2%) | 0.202 |
| Prior ischemic stroke or TIA, n (%) | 13 (12.4%) | 5 (5.2%) | 0.075 |
| Prior ICH, n (%) | 6 (5.7%) | 6 (6.3%) | 0.873 |
| Antiplatelet use, n (%) | 32 (30.5%) | 12 (12.5%) | 0.002 |
| **Clinical features** |  |  |  |
| Baseline SBP (mmHg), median (IQR) | 166 (151-186) | 160 (149-177) | 0.206 |
| Baseline DBP (mmHg), median (IQR) | 90.5 (78-105) | 90 (80-100.5) | 0.942 |
| Baseline NIHSS, median (IQR) | 16 (10-20) | 12 (7-16) | 0.002 |
| Baseline GCS, median (IQR) | 14 (12-15) | 15 (14-15) | 0.001 |
| Onset to scan time (mins), median (IQR) | 74 (54-87) | 74 (57-87) | 0.755 |
| Randomised to tranexamic acid, n (%) | 56 (50.5%) | 49 (51.0%) | 0.876 |
| **Baseline imaging features** |  |  |  |
| ICH location |  |  | 0.012 |
| Lobar, n (%) | 25 (23.8%) | 10 (10.4%) |  |
| Deep, n (%) | 80 (76.2%) | 86 (89.6%) |  |
| Presence of IVH, n (%) | 28 (26.7%) | 20 (20.8%) | 0.377 |
| Baseline ICH volume (ml), median (IQR) | 12.4 (5.7-23.6) | 8.1 (4.3-17.1) | 0.033 |
| Baseline IVH volume (ml), median (IQR) | 0 (0-0) | 0 (0-0.7) | 0.107 |
| Baseline hematoma growth rate (ml/hr), median (IQR) | 10.3 (4.3-26.8) | 7.3 (3.8-15.5) | 0.033 |
| **Outcomes** |  |  |  |
| 24-hour ICH volume (ml), median (IQR) | 15.3 (6.8-33.6) | 10.3 (5.5-21.7) | 0.110 |
| 24-hour IVH volume (ml), median (IQR) | 0 (0-3.3) | 0 (0-1.5) | 0.307 |
| Hematoma expansion at 24 hours, n (%) | 28 (32.6%; n=86) | 37 (40.2%; n=92) | 0.289 |
| 90-day mRS, median (IQR) | 4 (3-5) | 3 (2-4) | 0.002 |
| 90-day mortality, n (%) | 22 (21.0%) | 12 (12.5%) | 0.110 |

IQR = interquartile range; mRS = modified Rankin Scale; TIA = transient ischemic attack; ICH = intracerebral hemorrhage; SBP = systolic blood pressure; DBP = diastolic blood pressure; NIHSS = National Institute of Health Stroke Score; GCS = Glasgow Coma Scale; IVH = intraventricular hemorrhage

**Table S2:** Univariate logistic regression for ultra-early hematoma expansion and association with outcomes excluding patients with early deterioration (n=96)

|  | Unadjusted odds ratio (95% CI) | P-value |
| --- | --- | --- |
| 24-hour hematoma expansion | 38.33 (9.63-152.60) | <0.001 |
| Interval hematoma expansion* | 5.07 (1.33-19.24) | 0.017 |
| 90-day poor outcome (mRS 3-6) | 3.37 (1.04-10.89) | 0.042 |
| 90-day mortality | 8.55 (2.82-26.86) | <0.001 |

\*Hematoma expansion between 1-hour and 24-hour scans. mRS = modified Rankin Scale

**Supplementary Table S3:** Characteristics of participants without ultra-early hematoma expansion with 24-hour imaging (n=61)

|  | With 24-hour hematoma expansion (n=10) | Without 24-hour hematoma expansion (n=51) | p-value |
| --- | --- | --- | --- |
| **Demographics** |  |  |  |
| Age, median (IQR) | 72.5 (67-77) | 67 (53-75) | 0.203 |
| Female gender, n (%) | 2 (20%) | 19 | 0.295 |
| Region |  |  | 0.836 |
| Australia/New Zealand, n (%) | 10 (100%) | 50 (98%) |  |
| Asia, n (%) | 0 (0%) | 1 (2%) |  |
| Finland, n (%) | 0 (0%) | 0 (0%) |  |
| Mobile stroke unit recruitment, n (%) | 5 (50%) | 15 | 0.205 |
| Baseline mRS, median (IQR) | 0 (0-2) | 0 (0-0) | 0.554 |
| Past medical history |  |  |  |
| Hypertension, n (%) | 4 (40%) | 34 (66.7%) | 0.112 |
| Diabetes mellitus, n (%) | 2 (20%) | 7 (13.7%) | 0.609 |
| Atrial fibrillation, n (%) | 1 (10%) | 1 (2%) | 0.192 |
| Prior ischemic stroke or TIA, n (%) | 3 (30%) | 4 (7.8%) | 0.044 |
| Prior ICH, n (%) | 0 (0%) | 2 (3.9%) | 0.524 |
| Antiplatelet use, n (%) | 6 (60%) | 10 (19.6%) | 0.008 |
| **Clinical features** |  |  |  |
| Baseline SBP (mmHg), median (IQR) | 159 (152-173) | 167 (156-185) | 0.369 |
| Baseline DBP (mmHg), median (IQR) | 81.5 (74-100) | 92 (78-105) | 0.350 |
| Baseline NIHSS, median (IQR) | 17.5 (17-19) | 12 (8-18) | 0.083 |
| Baseline GCS, median (IQR) | 14 (13-15) | 15 (13-15) | 0.410 |
| Onset to scan time (mins), median (IQR) | 76 (50-88) | 73 (59-88) | 0.844 |
| Randomised to tranexamic acid, n (%) | 5 (50%) | 25 (49%) | 0.955 |
| **Baseline imaging features** |  |  |  |
| ICH location |  |  | 0.580 |
| Lobar, n (%) | 2 (20%) | 9 (17.6%) |  |
| Deep, n (%) | 8 (80%) | 42 (82.3%) |  |
| Presence of IVH, n (%) | 1 (10%) | 16 (31.4%) | 0.159 |
| Baseline ICH volume (ml), median (IQR) | 20.6 (8.4-27.3) | 8.4 (3.7-16.2) | 0.039 |
| Baseline IVH volume (ml), median (IQR) | 0 (0-0) | 0 (0-3.2) | 0.230 |
| Baseline hematoma growth rate (ml/hr), median (IQR) | 15.2 (5.8-28.5) | 6.5 (2.8-14.4) | 0.068 |
| **1-hour imaging features** |  |  |  |
| 1-hour ICH volume (ml), median (IQR) | 26.0 (9.9-29.7) | 8.3 (3.7-16.5) | 0.022 |
| 1-hour IVH volume (ml), median (IQR) | 0 (0-0) | 0 (0-5.0) | 0.411 |
| Absolute ICH growth (ml), median (IQR) | 2.6 (1.1-4.9) | 0.1 (-0.2-1.2) | 0.005 |
| Percentage ICH growth (%), median (IQR) | 17.6 (8.8-21.7) | 1.9 (-2.9-10.1) | 0.003 |
| **Interval imaging features** |  |  |  |
| Absolute ICH growth (ml), median (IQR) | 4.2 (3.8-8.9) | 0.1 (-0.8-1.1) | <0.001 |
| Percentage ICH growth (%), median (IQR) | 19.2 (18.1-26.6) | 1.6 (-10-9.1) | <0.001 |
| **Outcomes** |  |  |  |
| 24-hour ICH volume (ml), median (IQR) | 32.5 (14.0-36) | 8.3 (3.8-16) | 0.002 |
| 24-hour IVH volume (ml), median (IQR) | 0 (0-0) | 0 (0-3.7) | 0.430 |
| 90-day mRS, median (IQR) | 4.5 (3-5) | 3 (2-4) | 0.037 |
| 90-day mortality, n (%) | 1 (10%) | 4 (7.8%) | 0.820 |

IQR = interquartile range; mRS = modified Rankin Scale; TIA = transient ischemic attack; ICH = intracerebral hemorrhage; SBP = systolic blood pressure; DBP = diastolic blood pressure; NIHSS = National Institute of Health Stroke Score; GCS = Glasgow Coma Scale; IVH = intraventricular hemorrhage

**Supplementary Table S4:** Baseline characteristics of participants with ultra-early scans by randomisation group.

|  | Tranexamic acid (n=56) | Placebo (n=49) | p-value |
| --- | --- | --- | --- |
| **Demographics** |  |  |  |
| Age, median (IQR) | 69 (58.5-77.5) | 70 (53-78) | 0.241 |
| Female gender, n (%) | 24 (42.9%) | 18 (36.7%) | 0.523 |
| Region |  |  | 0.747 |
| Australia/New Zealand, n (%) | 53 (94.6%) | 48 (98.0%) |  |
| Asia, n (%) | 2 (3.6%) | 0 (0%) |  |
| Finland, n (%) | 1 (1.8%) | 1 (2%) |  |
| Mobile stroke unit recruitment, n (%) | 22 (39.3%) | 12 (24.5%) | 0.106 |
| Baseline mRS, median (IQR) | 0 (0-0) | 0 (0-0) | 0.721 |
| **Past medical history** |  |  |  |
| Hypertension, n (%) | 32 (57.1%) | 30 (61.2%) | 0.671 |
| Diabetes mellitus, n (%) | 10 (17.6%) | 8 (16.3%) | 0.836 |
| Atrial fibrillation, n (%) | 1 (1.8%) | 1 (2%) | 0.924 |
| Prior ischemic stroke or TIA, n (%) | 8 (14.3%) | 5 (10.2%) | 0.526 |
| Prior ICH, n (%) | 3 (5.4%) | 3 (6.1%) | 0.866 |
| Antiplatelet use, n (%) | 16 (28.6%) | 16 (32.7%) | 0.650 |
| **Clinical features** |  |  |  |
| Baseline SBP (mmHg), median (IQR) | 168 (154-188) | 164 (150-183.5) | 0.376 |
| Baseline DBP (mmHg), median (IQR) | 91 (78-107) | 90 (78-102) | 0.946 |
| Baseline NIHSS, median (IQR) | 17 (11-20.5) | 13 (10-19) | 0.104 |
| Baseline GCS, median (IQR) | 14 (12-15) | 14 (13-15) | 0.832 |
| Onset to scan time (mins), median (IQR) | 75.5 (57-87) | 73 (51-85) | 0.592 |

IQR = interquartile range; mRS = modified Rankin Scale; TIA = transient ischemic attack; ICH = intracerebral hemorrhage; SBP = systolic blood pressure; DBP = diastolic blood pressure; NIHSS = National Institute of Health Stroke Score; GCS = Glasgow Coma Scale

**Supplementary Figure S1:**
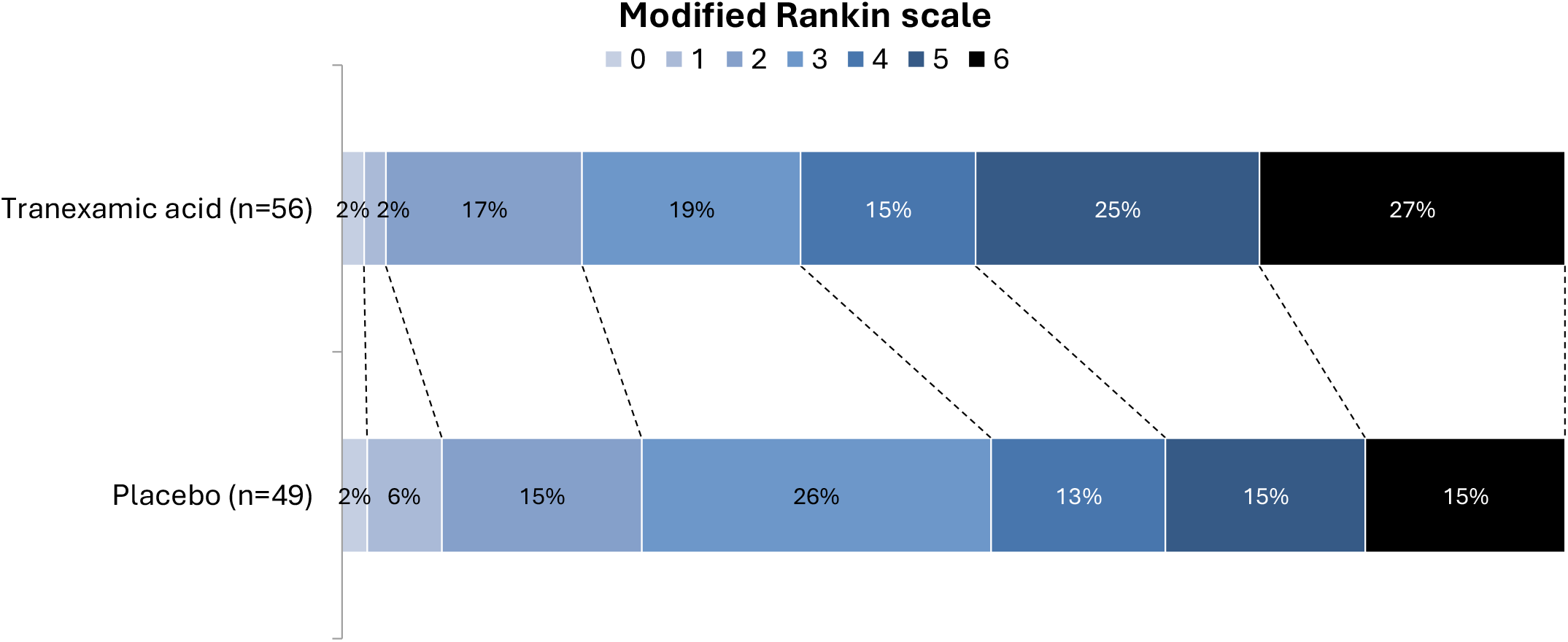
Modified Rankin Scale scores at 90 days for patients with 1-hour scans (unadjusted OR=0.55 [95% CI -0.13-1.23], p=0.115)

